# Examination of a novel limb symmetry index to discriminate movement strategies during bilateral jump landing in individuals with ACLR and with and without a history of ankle sprains

**DOI:** 10.1101/2024.03.18.24304515

**Authors:** Yuki A. Sugimoto, Craig J. Garrison, Ana Maria Acosta, Julius P.A. Dewald

## Abstract

**INTRODUCTION:** The Limb Symmetry Index (LSI), computed from kinetic parameters, tracks knee functionality post Anterior Cruciate Ligament Reconstruction (ACLR). However, LSI may lack accuracy in individuals with ACLR and ankle sprains, as it overlooks kinetic chain coordination across lower limb joints. Previous ankle sprains (AS) contribute to altered neuromuscular control in ACLR, emphasizing the need to evaluate within-limb coordination during bilateral tasks to prevent secondary ACL injuries. The effect of Energy Absorption Contribution (EAC) on joint work provides insight into the coordination between joints during observed movements. Thus, the purpose of this study was to validate a novel LSI based on EAC for discriminating movement strategies in bilateral drop vertical jump landing (DVJL) among individuals with ACLR and ACLR-AS

**METHODS:** 39 healthy athletes, including 13 healthy controls, 13 ACLR, and 13 ACLR-AS were matched by age, height, weight, sex, sports involvement, and limb dominance. Participants performed five DVJLs with kinematics and ground reaction forces recorded. Individual joint work (M) and EAC were calculated and averaged across the middle three trials to compute the LSI on individual joint work (LSI_M_) and EAC (LSI_EAC_). Negative LSI indicates asymmetry toward the nonsurgical limb, while positive LSI indicates asymmetry toward the surgical limb. A 3x2x3 repeated measures analysis of variance was utilized to analyze interactions between groups, the LSI method, and joint. Tukey’s LSD post-hoc analyses were used to examine within and between groups (alpha=0.05).

**RESULTS:** There was a significant interaction between the group, LSI method, and joint (F_4,72_=3.216, *P*=.017). LSI_EAC_ identified significant loading asymmetry at the hip (*P*=.046) and knee (*P*=.015) when compared to healthy controls.

**CONCLUSIONS:** LSI_EAC_ proved to be the method capable of distinguishing group differences in loading asymmetry at the hip and knee for the ACLR-AS group compared to healthy controls during bilateral DVJL. Overall, LSI_EAC_ provided a more comprehensive insight into the movement strategies employed during DVJL, particularly for individuals with ACLR and ACLR-AS. This emphasizes its suitability for effectively monitoring the rehabilitation progression post ACLR towards RTS. Our findings suggest that clinicians should prioritize computing the LSI based on EAC and assessing the history of ankle sprains for precise RTS decision-making. This approach not only aids in identifying loading asymmetry at the knee and hip but also underscores the critical importance of restoring modified movement strategies in adolescents with ACLR-AS, ultimately reducing the risk of secondary ACL injury following RTS.

## Background

Ankle sprains and anterior cruciate ligament (ACL) injuries are the most common sports-related musculoskeletal injuries. An estimated 23,000 ankle sprains occur each day, and up to 73%, of individuals with lateral ankle sprains experience repetitive ankle sprains, leading to the emergence of chronic ankle instability (CAI).^1–3^ Similarly, the annual incidence of ACL tears in the United States is reported to be 200,000 cases, with ACL reconstruction (ACLR) required in 90% of these cases.^4^ Notably, individuals who have undergone ACLR face a sixfold increased risk of experiencing secondary ACL injuries within the initial 24 months following their return-to-sports (RTS). The prevalence rate of a second ACL injury remains as high as 31%, particularly among adolescents.^4^

Previous research has suggested a potential link between CAI and ACLR.^5,6^ Indeed, 52% to 60% of individuals experiencing an ACL injury reportedly had prior encounters with ankle sprains.^5,6^ Anchored in the tenets of the lower extremity kinetic chain, individuals with CAI exhibit compensatory neuromuscular and biomechanical modifications at both the knee and the hip.^7–16^ During jump landing tasks, those with CAI demonstrated diminished knee flexion and increased hip flexion angles in the affected limb during jump landing tasks in comparison to healthy individuals and copers, who have a history of ankle sprains but did not develop CAI, respectively.^8,9^ Similarly, individuals with ACLR exhibit distinct landing patterns characterized by increased knee extension and reduced peak knee flexion moments, suggesting potential underloading of the surgical limb.^17^

Asymmetry in landing kinetics and kinematics, especially during a bilateral drop vertical jump landing, following ACLR, is identified as a significant risk factor for both initial and secondary ACL injuries.^4,18^ While much of the research focused on adults, a recent systematic review indicated that similar asymmetries exist in adolescents.^18,19^ These asymmetries include a reduced knee extension moment and vertical ground reaction force in the surgical limb during double-limb landing compared to the nonsurgical limb.^17,18^ Consequently, limb asymmetry index (LSI) is frequently utilized criterion, comparing the function and loading of the surgical limb to the nonsurgical limb, to make the decision to RTS.^20^ However, this assessment is primarily centered on the knee and overlooks the consideration of compensatory mechanisms within the kinetic chain at the ankle or hip. This oversight could potentially contribute to the occurrence of secondary ACL injuries, especially in ACLR individuals with a history of ankle sprains.

In the study conducted by Sigward et al. (2018)^21^, they examined the loading strategy employed during bilateral squatting at 3- and 5-months post-ACLR. The findings revealed that individuals with ACLR exhibited a combination of inter- and intra-limb compensations at 3 months.^21^ This involved shifting load from the surgical to the nonsurgical limb and redistributing load to the hip within the surgical limb.^21^ However, at 5 months post-ACLR, those with ACLR predominantly relied on intra-limb compensation, shifting load from the knee to the hip within the surgical limb.^21^ Robinson et al. (2022)^22^ identified a similar trend in adolescent athletes when investigating energy absorption contribution (EAC) at RTS. The EAC, evaluating the energy expended at each joint during a given task by combining both kinematic (joint angular velocity) and kinetic (net joint moment) data, was applied to scrutinize intra-limb loading across multiple joints (ankle, knee, hip). Notably, adolescent athletes with ACLR exhibited intra-limb compensation at the hip in the surgical limb by shifting the load from the knee.^22^ Additionally, there was intra-limb load-sharing compensation between the hip and knee and underloading of the ankle in the nonsurgical limb during DVJL.^22^

Our innovative approach, utilizing EAC instead of the conventional method of utilizing kinematic and kinetic data solely at the knee in the computation of LSI during DVJL, provides enriched insights by comprehensively considering both inter- and intra-limb compensations. This method has the potential to enhance the decision-making process for RTS, thereby reducing the risk of secondary ACL injuries. Consequently, the purpose of this study was to examine the efficacy of our novel approach in analyzing joint loading asymmetry based on EAC compared to the conventional approach in distinguishing loading asymmetry during DVJL in adolescent athletes with ACLR, with and without a history of ankle sprains, compared to healthy controls.

## Methods

### Participants

A total of thirty-nine athletes were recruited, comprising three groups: 13 healthy controls (15.08±1.38 years, 63.29±6.18 kg, 167.93±10.11 cm), ACLR (15.62±1.61 years, 69.40±13.94 kg, 167.28±7.16 cm), ACLR with a history of ankle sprains [ACLR-AS] (15.31±2.33 years, 74.30±14.34 kg, 170.13±9.50 cm) [Tbale 1]. Eligible participants, aged between 13 and 25 years, were enlisted based on their involvement in either a level 1 (e.g., basketball, football, soccer) or level 2 (softball, baseball) sport. For the ACLR and ACLR-AS groups, inclusion criteria required individuals to have sustained a first-time ACL injury, with specific exclusion criteria that excluded those with full-thickness chondral defects, grade II or III medial, lateral, or posterior collateral ligament injuries. Participants in the healthy control group were included if they had no active lower-extremity orthopedic injuries and had not experienced any injuries within the last three months.

All participants were carefully matched by age, weight, height, sex, sports involvement, and limb dominance in terms of ACLR. Participants’ surgical limb was defined as the limb with ACLR with and without a history of ankle sprains, and healthy controls were assigned a surgical limb based on their matched participant. If the participant’s surgical limb was dominant limb, the dominant limb (the limb participants would choose to kick a ball with) of healthy individuals was identified as the surgical limb and vice versa for the nonsurgical limb and nondominant limb.

Upon enrollment, each participant completed a demographic information sheet included details about injury history and sports participation. Informed consent, and for minors, parental consent along with child assent, were obtained from all participants. Additionally, all participants were part of a larger ongoing study examining clinical outcomes across the continuum of care, investigating movement profiles.

### Instruments (motion analysis)

A 10-camera Qualisys Motion Capture System (Qualisys AB, Göteborg, Sweden) with a capture rate of 120 Hz was used to capture joint kinematics in all three cardinal planes during the DVJL task. Thirty-three reflective markers were placed to each participant’s skin and clothing using double-sided tape to track bony landmarks, including spinous process of the seventh cervical vertebra, twelfth thoracic vertebra, between the fourth and fifth lumbar vertebrae, sternum, bilateral acromion process, anterior superior iliac spine, posterior superior iliac spine, greater trochanter, anterior mid-thigh, medial and lateral femoral epicondyles, anterior mid-shank, medial and lateral malleoli, calcaneus, and first and fifth metatarsal heads. Three additional retroreflective markers were attached on the sacrum as a cluster. Two AMIT (Advanced Mechanical Technology Inc., Watertown, MA) force plates, operating at 1200 Hz, were used to acquire joint kinetics.

### Drop Vertical Jump Landing

Participants underwent three trials of a drop vertical jump-landing (DVJL) task, following the protocol outlined by DiStefano et al. (2015, 2018)^23,24^ and Ward et al. (2018)^25^. The initiation involved standing on a box positioned at 50% of their height from the force plate. The JL task comprised two double-leg jumps: a forward jump from a 30 cm box onto both legs, followed immediately by a vertical jump for maximal height, as detailed by Padua et al. (2009)^26^. Trials where participants failed to reach the force plate or executed the take-off with a single foot were excluded from the analysis.

### Isokinetic Testing

The Biodex Multi-Joint Testing and Rehabilitation System (Biodex Medical Systems, Shirley, NY) was employed for isokinetic testing to assess extensor peak torque, referred to as quadriceps strength. The testing velocity was set at 60 degrees per second (60 deg/s). Participants were seated on the Biodex system and securely fastened with padded straps around the thigh, pelvis, and torso. This strapping aimed to minimize accessory and compensatory movements during testing (7,20,21,22). The alignment of the femoral condyle of the test limb followed the Biodex axis of rotation, as per the manufacturer’s instructions. To familiarize participants with the testing motion, five submaximal knee extension/flexion repetitions were performed. Subsequently, participants executed five consecutive concentric contractions to measure quadriceps strength at 60 deg/s. Testing commenced on the nonsurgical limb, followed by the surgical limb. For consistency, extensor peak torque was referred to as quadriceps strength. The mean of the five trials for each limb was normalized to body weight for subsequent data analysis.

### Data Processing and Reduction

For kinematic and kinetic data processing, Qualisys system-exported data were processed in Visual 3D, involving filtering with a Butterworth filter (12 Hz cutoff frequency). Lower extremity joint angles were determined via an inverse kinematic approach, and lower extremity internal moment was calculated with an inverse dynamic approach. Knee joint Energy Absorption was calculated by integrating the negative portion of the scalar power curve, normalized to height and weight and averaged across three trials. Joint EAC at each joint (ankle, knee, hip) was computed as a percentage of the total limb energy absorption (equation 1). The average of individual joint work and Energy Absorption Coefficient (EAC) was calculated across the central three trials to determine the LSI for both joint work (Equation 2) and EAC (Equation 3). A negative LSI denotes asymmetry favoring the surgical limb, while a positive LSI signifies asymmetry favoring the nonsurgical limb.

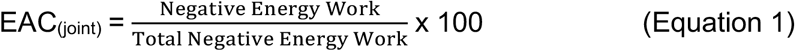

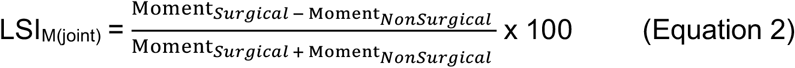

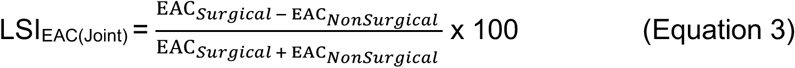

### Statistical Analysis

All statistical analyses were conducted using SPSS (version 27; IBM Corp, Armnok, NY, USA). A one-way analysis of variance was examined to compare participant characteristics (age, weight, weight, quads strength on the surgical and the nonsurgical limbs) among the groups (healthy controls, ACLR, ACLR-AS). Before conducting statistical analyses, normality and outliers in the kinetic data were examined. A 3 (group) × 2 (LSI methods) × 3 (joints) mixed-design repeated-measures analysis of variance was employed, with an alpha level set at 0.05, to examine how computing LSI with different methods at individual joints (ankle, knee, hip) differs among the three groups. Tukey’s LSD post-hoc analyses were used in the case of a significant interaction. In the presence of significant post-hoc pairwise comparisons, Cohen’s *d* effect size (ES) values were calculated, with categories defined as weak (≤0.40), moderate (0.40–0.80), and strong (≥0.80), along with corresponding 95% confidence intervals between the group means to assess the magnitude of difference in LSI methods and joints.^27^

## Results

Table 1 displays an overview of the participant characteristics within each group. There was a significant interaction between the group, LSI method, and joint (F_4,72_=3.216, *P*=.017). LSI_EAC_ identified significant loading asymmetry at the hip (*P*=.046) and knee (*P*=.015) when compared to healthy controls (Table 2).

**Table 1:** Participants’ demographics (Mean±SD)

|  | Healthy Controls | ACLR | ACLR-AS | *P*-Value |
| --- | --- | --- | --- | --- |
| N = 39 | 13 | 13 | 13 | - |
| Gender (Male; Female) | 5M; 8F | 5M; 8F | 5M; 8F | - |
| Age (years) | 15.08 ± 1.38 | 15.62 ± 1.61 | 15.31 ± 2.33 | *P* > .05 |
| Weight (kg) | 63.29 ± 6.18 | 69.40 ± 13.94 | 74.30 ± 14.34 | *P* > .05 |
| Height (cm) | 167.93 ± 10.11 | 167.28 ± 7.16 | 170.13 ± 9.50 | *P* > .05 |
| ACLR on the Dominant-Limb | 6 | 6 | 6 | - |
| ACLR on the NonDominant-Limb | 7 | 7 | 7 | - |
| Quads Strength (Nm/kg): Surgical Limb | 1.69 ± .52 | 1.59 ± .36 | 1.80 ± .62 | *P* > .05 |
| Quads Strength (Nm/kg): NonSurgical Limb | 1.99 ± .60 | 1.91 ± .41 | 2.00 ± .41 | *P* > .05 |

Abbreviations: ACLR, Anterior Cruciate Ligament Reconstruction; Quads, Quadriceps; M, Male; F, Female; ACLR-AS, Anterior Cruciate Ligament Reconstruction With A History of Ankle Sprains.

**Table 2:** A 3-Factor Interaction for Group, LSI method, and joint: Between Group Pairwise Comparisons for joint loading asymmetry (Mean±SD).

| Parameter |  | Group |  |  | P-Value | Effect Sizes (95% CI) |
| --- | --- | --- | --- | --- | --- | --- |
| LSI Method | Joint | Healthy Controls | ACLR | ACLR-AS |  |  |
| Joint Moment | Hip | -62.96 ± 49.85 | -47.07 ± 16.59 | | 0.229 | 0.43 (-0.17 to 1.03) |
| | | -62.96 ± 49.85 | | -58.98 ± 22.84 | 0.760 | 0.10 (-0.49 to 0.69) |
| | | | -47.07 ± 16.59 | -58.98 ± 22.84 | 0.365 | 0.60 (-0.01 to 1.20) |
| | Knee | -28.60 ± 44.35 | -13.30 ± 36.08 | | 0.354 | 0.38 (-0.22 to 0.97) |
| | | -28.60 ± 44.35 | | -37.03 ± 43.66 | 0.608 | 0.19 (-0.40 to 0.78) |
| | | | -13.30 ± 36.08 | -37.03 ± 43.66 | 0.154 | 0.59 (-0.01 to 1.20) |
| | Ankle | -84.40 ± 55.41 | -92.39 ± 3.58 | | 0.451 | 0.20 (-0.39 to 0.80) |
| | | -84.40 ± 55.41 | | 94.01 ± 4.08 | 0.530 | 0.24 (-0.35 to 0.84) |
| | | | -92.39 ± 3.58 | 94.01 ± 4.08 | 0.899 | 0.42 (-0.18 to 1.02) |
| EAC | Hip | 63.04 ± 12.78 | 54.33 ± 17.15 | | 0.184 | 0.58 (-0.03 to 1.18) |
| | | 63.04 ± 12.78 | | 49.74 ± 18.70 | **0.046\*** | 0.83 (0.21 to 1.45) |
| | | | 54.33 ± 17.15 | 49.74 ± 18.70 | 0.480 | 0.30 (-0.34 to 0.85) |
| | Knee | 11.39 ± 7.67 | 16.20 ± 17.17 | | 0.409 | 0.36 (-0.23 to 0.96) |
| | | 11.39 ± 7.67 | | 26.15 ± 17.08 | **0.015\*** | 1.11 (0.48 to 1.75) |
| | | | 16.20 ± 17.17 | 26.15 ± 17.08 | 0.092 | 0.58 (-0.02 to 1.18) |
| | Ankle | 25.57 ± 12.56 | 29.48 ± 9.86 | | 0.335 | 0.05 (-0.54 to 0.64) |
| | | 25.57 ± 12.56 | | 24.12 ± 7.56 | 0.719 | 0.11 (-0.48 to 0.70) |
| | | | 29.48 ± 9.86 | 24.12 ± 7.56 | 0.189 | 0.06 (-0.54 to 0.65) |

Abbreviations: LSI, Limb Symmetry Index; ACLR, Anterior Cruciate Ligament Reconstruction; ACLR-AS, Anterior Cruciate Ligament Reconstruction With A History of Ankle Sprains; CI, Confidence Intervals; EAC, Energy Absorption Contribution.
\*Indicates a significant difference.

## Discussion

The purpose of this study was to examine the effectiveness of our novel approach in contrast to traditional methods utilizing joint moment to discriminate joint loading asymmetry during DVJL among adolescent athletes with ACLR and ACLR-AS compared to healthy controls. The most important finding of the study was that group differences in joint loading asymmetry depended on both joint (hip, knee, ankle) and LSI computational methods (LSI_EAC_, LSI_M_). The pairwise comparisons demonstrated that only LSI_EAC_ effectively identified group differences in joint loading asymmetry at the hip and knee in adolescent athletes with ACLR-AS compared to healthy controls at RTS. Additionally, there was a significant directional loading asymmetry at individual joints when comparing LSI_EAC_ and LSI_M_ within each group, with LSI_EAC_ revealing joint loading compensation in the surgical limb, while LSI_M_ exhibited joint loading compensation in the nonsurgical limb. These collective findings indicate that LSI_EAC_ accurately discriminates loading asymmetry at the hip and knee in adolescent athletes with ACLR-AS at RTS.

To the best of our knowledge, this is the first study that compares a novel exploration of loading asymmetry and a conventional approach in ACLR adolescents with and without a history of ankle sprains. Prior investigations have predominantly focused on knee kinetics and kinematics during bilateral DVJL at RTS.^18^ These studies consistently identified limb asymmetry in parameters such as peak knee extension moment, peak vertical ground reaction force (vGRF), knee energy absorption, and peak knee flexion in the nonsurgical limb.^17,28–30^ Adolescents with ACLR employ a compensatory strategy by underloading the surgical limb compared to the nonsurgical limb.^18^ While the current study did not conduct direct within-group comparisons for limb asymmetry, the directional limb asymmetry identified through LSI_M_ aligns with previous research. Specifically, across all groups, there was an almost consistent strategy of underloading the surgical limb in the ACLR and ACLR-AS groups, as well as underloading an assigned surgical limb (nondominant limb) in healthy controls.

There were a limited number of prior studies examining limb asymmetry, including healthy controls, and they did not explicitly investigate group differences compared to the ACLR group. Paterno et al. (2011)^31^ found no limb asymmetry in peak vGRF, while Mueske et al. (2018)^32^ identified reduced knee flexion in the nonsurgical limb (nondominant limb) than in the surgical limb (dominant limb) in healthy controls. Limb dominance has been proposed to positively impact limb asymmetry of the knee extensor strength, indicating smaller strength deficits in those individuals who underwent ACLR in their dominant limb (Zumstien et al., 2022).^33^ In our study, both adolescent athletes with ACLR and ACLR-AS exhibited nearly equal numbers of adolescent athletes who sustained ACLR in their dominant limb (N=6) and those in their nondominant limb (N=7). Furthermore, there were no significant differences in quadriceps strength among all three groups in our study (Table 1). The conventional method for computing LSI doesn’t consider inter-limb compensation at the hip and ankle. Consequently, there is a potential for LSIM to overestimate joint function at RTS, thereby contributing to a secondary ACL injury.^30^ The collective absence of identified group differences in limb asymmetry at the hip, knee, and ankle may stem from movement strategies implemented in other joints of the kinetic chain that LSIM is not designed to detect.

Our novel approach, diverging from the conventional method to compute LSI (LSI_M_), involves computing LSI based on EAC (LSI_EAC_). Based on the work of Sigward et al., (2018)^21^ with the recognition that limb asymmetry is more prevalent in kinetic rather than kinematic variables among ACLR individuals, LSI_EAC_ entails the computation of EAC. This process utilizes both kinetic and kinematic data to assess the energy contributions from the hip, knee, and ankle during DVJL, emphasizing inter-limb compensation.^21^ Sigward et al. (2018)^21^ delved into the loading strategy adopted during bilateral squatting at 3- and 5-month intervals post-ACLR.^21^ The results highlighted a blend of inter- and intra-limb compensations at the 3-month mark among those who underwent ACLR, involving load transfer from the surgical to the nonsurgical limb and redistribution within the surgical limb.^21^ By the 5-month post-ACLR mark, a predominant shift to intra-limb compensation occurred, with the load shifting from the knee to the hip within the surgical limb.^21^ Accordingly, the application of EAC to compute LSI_EAC_ in our study unveiled limb asymmetry at the hip and knee in the ACLR-AS group, distinguishing them from both the ACLR group and healthy controls.

The absence of limb asymmetry at the ankle in adolescent athletes with ACLR-AS and the lack of limb asymmetry compared to healthy controls and the ACLR group were unexpected findings. Typically, limb asymmetry persists for approximately 8 to 12 months after ACLR but tends to diminish over time post-ACLR.^34^ At the time of RTS, individuals with ACLR with weaker quadriceps strength are known to exhibit altered landing patterns, whereas those with nearly symmetric quadriceps strength show landing patterns comparable to those of healthy controls during DVJL.^34^ In our study, all three groups demonstrated similar quadriceps strength at RTS, suggesting that the absence of strength deficits may not have contributed to differences in landing mechanics among individuals with ACLR (Table 1). However, it is important to note that distal changes caused by ankle sprains may have a significant impact on proximal joint kinematics in individuals with ACLR-AS.

Individuals experiencing multiple ankle sprains, such as CAI, demonstrate neuromuscular alterations at the knee and hip. Specifically, individuals with a history of ankle sprains show reduced knee flexion angles and increased hip flexion angles in the affected limb during jump landing tasks.^8,10^ Moreover, those with a history of ankle sprains had increased general joint laxity and genu recurvatum.^5,6^ The current evidence suggests there is a relationship between a history of prior ankle sprains and ACL injuries in athletes.^5,6^ Indeed, 52 to 60% of individuals who sustained an ACL injury had a prior ipsilateral ankle sprain.^5,6^ Earlier studies that observed limb asymmetry at RTS did not investigate the history of ankle sprains in their participants with ACLR. Therefore, it is plausible that those with ACLR displaying persistent limb asymmetry at the time of RTS had a history of ankle sprains, impacting their movement strategies and loading patterns compared to individuals with ACLR. Overall, LSI_EAC_ was sensitive enough to detect subtle differences in limb asymmetry, particularly in athletes with ACLR-AS.

### Clinical Implications

The risk of secondary ACL injury remains high, particularly among adolescents, with a prevalence rate reaching up to 31%.^4^ Limb asymmetry in landing kinetics and kinematics, particularly evident in a bilateral DVJL after ACLR, is acknowledged as a significant risk factor for secondary ACL injuries.^28^ This increased risk could be linked to overestimated limb asymmetry utilizing conventional LIS based on kinematic and kinetic data solely at the knee in the computation of LSI to make RTS decisions. Furthermore, despite up to 60% of individuals with ACLR reporting prior ankle sprains and the observed intra-limb compensation predominantly relied upon 5-months post-ACLR, the current RTS decision-making process tends to disregard compensatory mechanisms within the kinetic chain, particularly in individuals with ACLR and ACLR-AS.^5,6,21^ Based on our findings, clinicians should consider computing LSI based EAC and assessing the history of ankle sprains for accurate RTS decision-making. This approach not only facilitates the identification of limb asymmetry at the knee and hip but also emphasizes the importance of restoring modified movement strategies in adolescents with ACLR-AS. This emphasis is crucial for preventing the occurrence of secondary ACL injury following RTS.

### Limitations

The study sample includes adolescents and an athletic population, indicating possible constraints in extending these results to older or less active cohorts. Moreover, we lack information on the number of prior ankle sprains, the duration since the last ankle sprain, or any history of additional lower extremity musculoskeletal injuries that could potentially impact limb asymmetry during DVJL. Finally, we did not match subjects by graft type, limiting our ability to offer conclusive recommendations on the effects of graft type on limb asymmetry identified by LSI_EAC_.

## Conclusion

LSI_EAC_ proved to be the method capable of distinguishing group differences in loading asymmetry at the hip and knee for the ACLR-AS group compared to healthy controls during bilateral DVJL. Overall, LSI_EAC_ provided a more comprehensive insight into the movement strategies employed during DVJL, particularly for individuals with ACLR and ACLR-AS. This emphasizes its suitability for effectively monitoring the rehabilitation progression post ACLR towards RTS. Our findings suggest that clinicians should prioritize computing the LSI based on EAC and assessing the history of ankle sprains for precise RTS decision-making. This approach not only aids in identifying loading asymmetry at the knee and hip but also underscores the critical importance of restoring modified movement strategies in adolescents with ACLR-AS, ultimately reducing the risk of secondary ACL injury following RTS.

## Data Availability

All data produced in the present work are contained in the manuscript

## Acknowledgement

We would like to thank Shiho Goto, PhD, ATC, the Texas Health Sports Medicine physician, staff, and research coordinators for their valuable assistance in collecting data.

